# A comparison of pain registration by Visual Analog Scale and Numeric Rating Scale – a cross-sectional study of primary triage registration

**DOI:** 10.1101/2020.11.03.20225367

**Authors:** Jofrid Kollltveit, Malin Osaland, Marianne Reimers, Magnus Berle

## Abstract

**Background:** Pain is a subjective sensation; self-reporting is important for quantifying pain intensity. There are several different validated tools for this, such as Visual Analog Scale and Numeric Rating Scale. In the clinic, these terms are often used as equivalent. The objective of this study was to examine correlation and agreement between the pain registration tools in triage in an emergency department.

**Materials and Methods:** The study was performed in the Department of Emergency Medicine at Haukeland University Hospital in the period June-August 2019. We registered the pain score with two tools in 200 unselected patients in emergency admission with pain. In addition, we registrered gender, age, triage and general department affiliation.

**Results:** We found a strong correlation between the pain registration tools by Spearmans correlation test (rho=0,930, p<0,001). There were no significant difference between the pain registration tools within the subgroups. Bland-Altman analysis show agreement between the two pain registration tools.

**Conclusions:** In an Emergency Department triage is it acceptable to use Visual Analog Scale and Numeric Rating Scale as equivalent, as long as the correct terminology is used.

## Background

Pain is a subjective sensation, which is important to quantify in a clinical setting (1, 2). Pain score is a part of defining the degree of urgency of diagnostics and treatment, and is important for evaluating the effect of treatment. There are several validated tools to rate pain, as Visual Analog Scale (VAS) and Numeric Rating Scale (NRS) (2, 3). Using VAS, the patient defines the degree of pain on a visual scale without numeric values, while the values are visible on the back side for the registrar. NRS is a verbal numeric scale, where the patient grades their own pain on a scale between 0 and 10. Both methods are much used in clinical practice and research (3). In the clinic, the terms are often used as interchangeable, and NRS is in general noted as VAS (4). Several studies have shown a strong statistical correlation between VAS and NRS scores (1, 5, 6, 7). Some studies show good agreement between methods (1, 6) while others find a larger discrepancy (5, 7, 8). The objective of this study was to examine correlation and agreement between VAS and NRS score in patients admitted with pain to an emergency department in Norway.

## Materials and Methods

We performed a cross-sectional study of pain in primary triage registration at the emergency department at Haukeland University Hospital in the period June to August 2019. As a part of the primary triage at admission, the nurse collected pain score by use of NRS. The nurse asked the patient to grade their pain with an integer between 0 and 10, where 0 is no pain and 10 is the worst thinkable pain. After the nurse had finished the triage, the patients were asked to participate in a study. After oral approval, the patient was again scored with VAS. We used a horizontal VAS-ruler (0-100 mm) with the endpoints “no pain” to the left and “worst thinkable pain” to the right. The patient graded their pain with a moveable plastic marker to their pain level. We transcribed the value in mm and converted to the closest integer (0-10). The following variables were registered on paper: VAS, NRS, gender, age, triage and general department affiliation.

The target population were patients with pain in triage in the Emergency Department. We made a general estimation in advance on the number of patients to include, based on presumed sufficient material and a general achivable number. 200 unselected patients with pain were included, no clinical groups of patients were excluded. Exclusion criteria were age under 18 years, lack of competence to consent, cognitive failure as well as lack of lingual or motor skills.

The study included patients after informed oral consent. We have in our study design refrained from written informed consent to reduce the amount of registered personal information. The patient information was anonymized immediately after inclusion. The procedure of inclusion was made in agreement with the Data Protection Officer of the Hospital and the Regional Ethics Board of Western Norway.

We predefined a *null hypothesis* of no significant difference between VAS and NRS value. *A priori* acceptable clinical difference between the scores were defined to a maximum of +/-1, based on repeatability of test-retest of VAS (1). The collected data were organized in MS Excel, then analyzed in SPSS v2.5. We utilized Spearman correlation coefficient and Bland-Altman plot to study correlation and agreement between VAS and NRS value. Kruskal-Wallis test was used for the subgroup analyses between VAS and NRS for gender (female/male), age (over/under 30 years old), triage (green/yellow/orange) and general department affiliation (surgery/medicine)

## Results

Table 1 shows descriptive statistics with mean for VAS and NRS with standard deviation, as well as values for statistical tools. Spearman correlation coefficient (rho = 0,93) show strong correlation between VAS and NRS value (p < 0,001). Subgroup analysis by Kruskal-Wallis test does not show significant differences between VAS and NRS-value within the subgroups (p < 0,05). Bland-Altman plot (Figure 1) show agreement between VAS and NRS-value as pain scores.

**Table 1:** Descriptive statistics and statistical calcaltions

**Figure 1:**
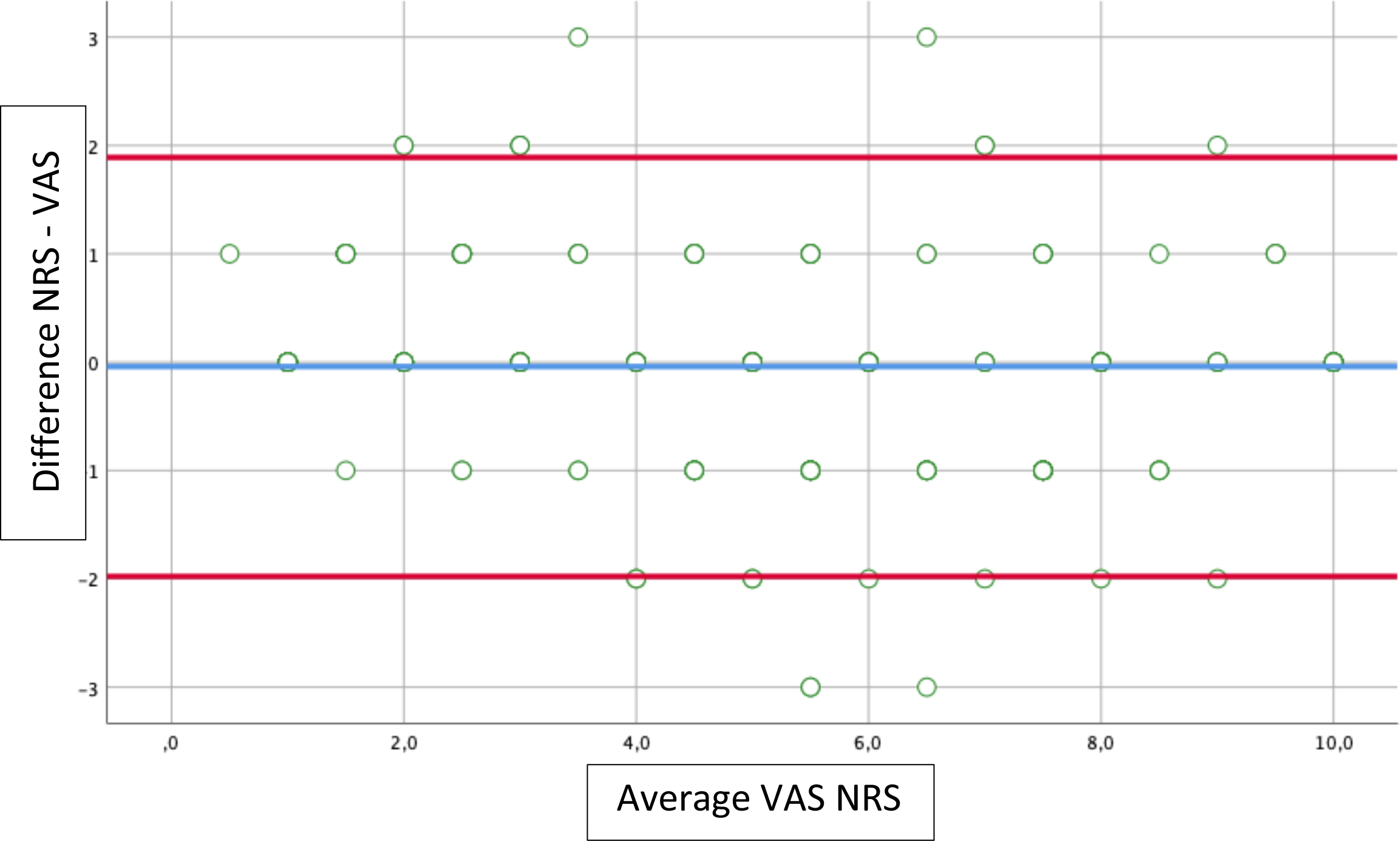
Bland-Altman plot of NRS vs VAS.

## Discussion

The strength of this study is that pain was reported by VAS short time after NRS by standardized questions in such a way that the information to the patient group was the same. The data collection was in daytime and evening in an un-selected patient group to ensure a more representative material.

The study has some limitations. Presenting VAS short time after NRS may introduce a bias. Collection of oral informed consent creates an emphasis on the fact that this is a study, with risk of patients attempting to answer the same. All patients were presented to NRS before VAS, where the triage nurse collected NRS-value while the VAS-value was collected by two medical students. We do only present a single time point and the data cannot tell of changes in VAS versus NRS over time. We have not examined which method is the better, only whether the two methods produce equal results. Earlier studies establish a consensus that NRS is easier to perform than VAS (2, 9). The results of this study do not establish that the method is equal in all populations. There are similar studies in other populations, among them orthopedic patients and cancer patients. These studies show good correlation between VAS and NRS (10, 11).

There are conflicting studies on the agreement between NRS and VAS (1, 5, 6, 7, 8). Our results support the studies showing good correlation and agreement between the methods (1, 6). We cannot abandon the null hypothesis of no significant difference and the limits of agreement in Bland-Altman plot is whitin the a priori clinical acceptable difference. This can indicate that the methods are interchangeable. NRS being noted as VAS will not cause errors in the clinic but we would advocate the use of proper terminology.

## Conclusions

The study shows a strong statistical correlation and a good agreement for reported scores between VAS and NRS. In an Emergency Department triage is it acceptable to use Visual Analog Scale and Numeric Rating Scale as equivalent, as long as the correct terminology is used.

## Supporting information

Anonymized registered data

Descriptive statistics and statistical calcaltions

## Data Availability

The data in anonymized form is included as supplementary data.

## List of abbreviations

VAS: Visual Analog Scale
NRS: Numeric Reported Scale

## Declarations

### Ethics approval and consent to participate

The study was approved by the Data Protection Officer of the Hospital and the Regional Ethics Board of Western Norway, approval REK Vest (2019/484).

### Consent for publication

Patients were included by oral consent to limit registered personal information in accordance with the Data Protection Officer and the Regional Ethics Committee.

### Availability of Data

The data in anonymized form is included as supplementary data.

### Authors’ contributions

Patient inclusion and data collection JK, MO. Project plan, implementation and data analysis JK, MO, MR, MB. Writing of manuscript JK, MO, MB. Original idea MB. All Authors read and approved the final manuscript.

## Figure and table text

Additional file 1: Anonymized registered data

